# Novel Hypodensity Detection Tool improves clinician identification of hypodensity on non-contrast CT in stroke patients

**DOI:** 10.1101/2023.11.02.23298012

**Authors:** A Dos Santos, M Visser, L Lin, A Bivard, L Churilov, M Parsons

**Affiliations:** University of New South Wales, South-Western Sydney Clinical Campus, NSW, Australia. University of Melbourne, VIC, Australia; University of Melbourne, Melbourne Brain Centre, Melbourne, VIC, Australia; Sydney Brain Centre, Faculty of Medicine, University of New South Wales, NSW, Australia; Melbourne Medical School, University of Melbourne, Melbourne, VIC, Australia; University of New South Wales, South-Western Sydney Clinical Campus, NSW, Australia. Department of Neurology, Liverpool Hospital. Ingham Institute for Applied Medical Research Liverpool, NSW, Australia

## Abstract

**Background:** In acute stroke, identifying early changes (parenchymal hypodensity) on non-contrast CT (NCCT) can be challenging. We aimed to identify whether the accuracy of clinicians in detecting acute hypodensity in ischaemic stroke patients on a non-contrast CT is improved with the use of an automated HDT algorithm using MRI-DWI as the gold standard.

**Methods:** The study employed a case-crossover within-clinician design, where clinicians were tasked with identifying hypodensity lesions on NCCT scans for five a priori selected patient cases, before and after viewing the HDT. The DICE Similarity Coefficient (DICE score) was the primary measure of accuracy. Statistical analysis compared DICE scores with and without HDT using mixed-effects linear regression, with individual NCCT scans and clinicians as nested random effects.

**Results:** The HDT had a mean DICE score of 0.62 for detecting hypodensity across all NCCT scans and clinicians overall mean DICE score of 0.33 (SD 0.31) before HDT implementation and 0.40 (SD 0.27) after implementation. HDT use was associated with an increase of 0.07 (95% CI: 0.02-0.11, p=0.003) in DICE score accounting for individual scan and clinician effects. For scans with small lesions, clinicians achieved a mean increase in DICE score of 0.08 (95% CI: 0.02, 0.13, p=0.004) following HDT use. In a subgroup of 15 trainees, DICE score improved with HDT implementation (mean difference in DICE 0.09 [95% CI: 0.03, 0.14, p=0.004]).

**Conclusions:** The Hypodensity Detection Tool (HDT) has potential to enhance accuracy of detecting hypodensity in acute stroke diagnosis, especially for smaller lesions, and notably for less experienced clinicians.

## Introduction

The mainstay of acute stroke imaging has been Computed Tomography (CT). It is relatively accessible in most hospitals throughout the world, is inexpensive compared with Magnetic Resonance Imaging (MRI), efficient, fast and has few contraindications.(1) However, in the first few hours after stroke onset, identification of the early signs of ischaemic stroke (parenchymal hypodensity and focal swelling) on non-contrast CT (NCCT)(1-3) can be challenging, for even the most experienced clinicians.(4) Image interpretation can delay therapeutic decisions and is often the rate limiting step, particularly if the radiologist is offsite, which often is the case in rural and remote Australia for example.(5, 6) For the onsite clinicians, fatigue and inexperience can affect image interpretation and delay treatment decisions.(1) However, identification of these subtle changes (particularly parenchymal hypodensity) is necessary as they likely represent irreversible ischaemia and this is an important consideration in the decision to offer reperfusion therapy.(6)

As reperfusion treatment is time critical, decision support tools such as automated hypodensity detection have the potential to improve detection of early ischemic change and reduce delays in diagnosis and reperfusion treatment.(1)

Current literature describes several approaches for hypodensity detection. For example, image filtering (windowing) to enhance the visibility of ischaemic changes(7, 8), spatial normalisation between a template of healthy controls and the examined brain(9, 10), topographic scoring using the territories of the middle cerebral artery (MCA)(11, 12), classification of the image texture features(13) and imaging biomarkers(14).

One automated NCCT hypodensity detection tool (HDT, MIstar, Apollo Medical Imaging, Melbourne, Australia) uses histogram-based left-right brain comparisons to detect regions-of-interest that show unilateral hypodense areas. It uses iterative level-set optimization to identify areas of hypodensity within a non-contrast CT scan. This HDT showed strong positive correlation with the gold standard, Magnetic Resonance Imaging Diffusion-weighted Imaging (MRI-DWI) (correlation coefficient>0.5, unpublished data).

Thus, the aim of this study was to investigate whether the accuracy of clinicians in detecting acute hypodensity in ischaemic stroke patients on non-contrast CT is improved with the use of an automated HDT algorithm using MRI-DWI as the gold standard.

## Methods

### Study design

This was a case-cross over within-clinician study where clinicians were asked to identify hypodensity lesions first before, and then after, the help of the HDT output on five a priori purposively sampled patient cases that represent the broad population of acute stroke patients. Thirty-two clinicians were approached to participate in the study. The clinicians were either neurology trainees with less than 5 years experience reading NCCT in acute stroke patients or consultant neurologists with more than 5 years experience. Recruiting 32 clinicians provided 0.8 power to detect the medium effect size (Cohen’s d=0.5) for the difference in means of DICE score (dependent samples/matched pairs without and with the use of HDT) under the settings of two-sided Type I error of 0.05.

Clinicians were instructed to manually segment a single slice of five different patients’ NCCT scans before having access to the HDT output. Once they had segmented the single slice and sent their segmentation back to the study coordinator (MV), they were given the HDT output that had identified the hypodensity on the same single slice. The clinicians were asked to compare the HDT output and their initial drawing. They were asked to draw a new lesion if they felt the HDT helped identify the hypodensity. They then sent their completed segmentations back to the study coordinator for analysis.

### Study scans

Five NCCT scans with a corresponding single slice, from 2 sites (John Hunter Hospital and Royal Adelaide Hospital) were selected from the International Stroke Perfusion Registry (INSPIRE) using the following inclusion criteria: 1) high-resolution NCCT at admission, 2) occlusion of the anterior circulation and 3) MRI-DWI obtained during admission.

The 5 scans were purposively selected to represent variant lesion size and hemisphere, representative of acute stroke, of the anterior circulation. Among the 5 NCCT scans, 2 had large lesions with hypodensity involving more than 1/3 the MCA territory, and 3 scans had small lesions.

### Automated NCCT hypodensity detection tool

The NCCT scans in this study were obtained from the Toshiba Aquilion One (Canon, Tokyo, Japan). NCCT lesion was segmented automatically with the HDT algorithm on MIStar software as illustrated in Figure 1 (Apollo Medical Imaging, Melbourne, Australia). It consisted of the following steps: 1) assessing the symmetry of the density histograms of the left and right hemisphere (after registration to a template), 2) definition of potential seeds, and 3) iterative optimization of level-set thresholds.

**Figure 1.**
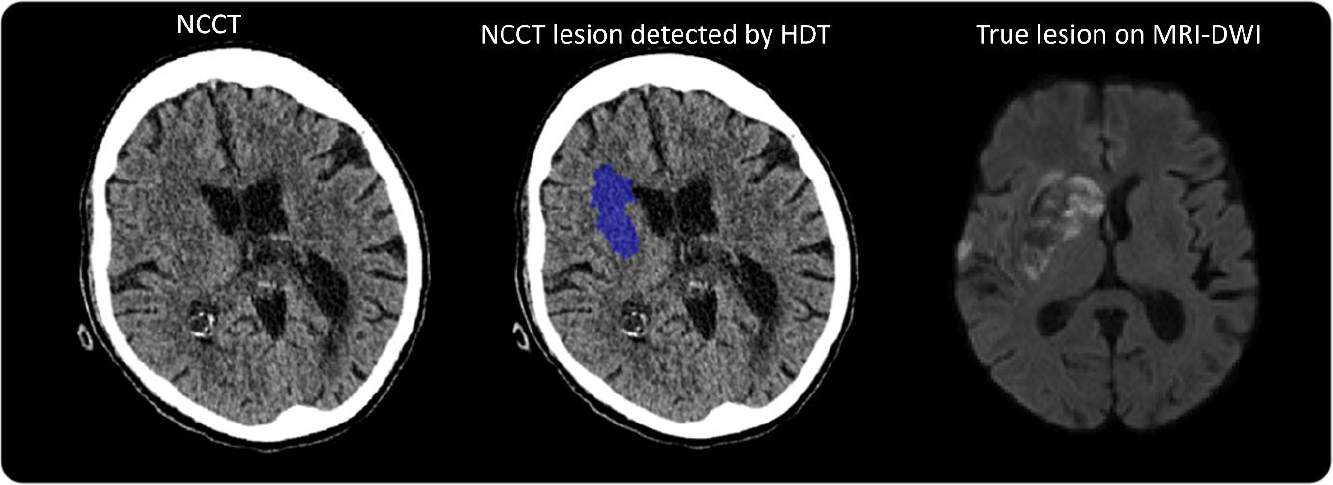
NCCT hypodensity detection tool (HDT) example.

### Manual segmentation of NCCT

The clinicians were instructed to manually segment on axial views, a slice of each of the five NCCT scans. Clinicians were provided the whole brain NCCT for review only. The segmentation was completed using the paintbrush mode in the ITK-SNAP software application (http://www.itksnap.org).

### Manual segmentation of MRI

MRI-DWI images were manually segmented by trained personnel using ITK-SNAP to extract infarct lesions as reference. MRI-DWI lesions were registered to the NCCT images using Advanced Normalization Tools (ANTS).(15)

### Statistical analysis

The primary outcome of this study was DICE Similarity Coefficient (DICE score). The DICE score measures the similarity of the lesion segmentations on NCCT and on MRI-DWI. It ranges from 0 to 1, when 0 represents no overlap and 1 represents perfect overlap. The DICE score is calculated by the following equation:

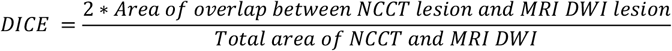

In this study, the DICE scores were calculated for the following comparisons: 1) the HDT output and the registered MRI-DWI lesion, 2) the clinicians’ segmentation before viewing the HDT and the registered MRI-DWI lesion, and 3) the clinicians’ segmentation after viewing the HDT and the registered MRI-DWI lesion.

The DICE scores were summarized using mean and standard deviation (SD). To compare the difference of the DICE score for clinicians before and after HDT implementation, a three-level mixed-effects linear regression was performed with DICE score as the outcome, HDT implementation before versus after as the independent variable, and NCCT scans and clinicians as nested random effects.

Subgroup analyses were conducted on NCCT slices with large and small lesions, as well as NCCT slices segmented by consultants versus trainees.

All statistical analyses were performed with STATA 13.0 (Stata Corp, College Station, Texas, USA). p-values less than 0.05 were considered as indicative of statistical significance. Confidence intervals (CI) were set at 95%.

## Ethics

This study used data from the INSPIRE registry. The INSPIRE had central ethics approval by the Hunter New England Human Research Ethics Committee (HNEHREC 11/08/17/14.01), Written informed consent was obtained for each patient for their data to be used as part of the INSPIRE registry. The INSPIRE registry and all associated analyses are conducted in accordance with the declaration of Helsinki.

## Results

### Data Selection

A total of 32 clinicians participated in the study. Five clinicians were excluded as their drawings were incorrectly saved and unable to be read. For the remaining 27 clinicians, 26 clinicians had the 5 NCCT scan assessments completed before and after the HDT was provided, whereas 1 clinician had 2 NCCT scans reviewed and assessed. Thus, a total of 132 matched pairs of segmentations before/after HDT from 27 clinicians were included in this study.

### HDT performance against MRI-DWI gold standard

When compared to the true lesion reference on MRI-DWI, the HDT resulted in a mean DICE score of 0.62 (SD 0.05) in detecting the hypodensity region on NCCT slices. The HDT performance was consistent across the 5 NCCT slices, with DICE scores ranging from 0.54 to 0.66 (Table 1).

**Table 1:** Median DICE score across the 6 NCCT for clinicians and HDT.

| Image ID | N | Clinician DICE before HDT implementation, mean (SD) | Clinician DICE after HDT implementation, mean (SD) | DICE of HDT |
| --- | --- | --- | --- | --- |
| 1 | 26 | 0.08 (0.18) | 0.17 (0.22) | 0.54 |
| 2 | 27 | 0.30 (0.32) | 0.41 (0.28) | 0.63 |
| 3 | 26 | 0.58 (0.21) | 0.55 (0.20) | 0.62 |
| 4 | 27 | 0.27 (0.32) | 0.35 (0.30) | 0.66 |
| 5 | 26 | 0.42 (0.24) | 0.53 (0.17) | 0.65 |
| Overall | 132 | 0.33 (0.31) | 0.40 (0.27) | 0.62 |

### Clinician performance against MRI-DWI gold standard

The overall mean DICE score for clinician interpretation of the NCCT slice before the HDT was applied was 0.33 (SD 0.31). The mean DICE score was 0.40 (SD 0.27) after clinicians viewed the HDT output (Table 1).

HDT use was associated with an increase of 0.07 (95% CI: 0.02-0.11, p=0.003) in mean DICE score on mixed-effects linear regression, accounting for individual scans and clinicians as nested random effects.

For each NCCT, the performance of clinicians with and without the HDT in delineating NCCT lesion is summarized in Table 1, Table 2, and Figure 2. The mixed-effects linear regression showed that DICE score was significantly increased in 4 NCCT scans after the HDT was implemented, with the mean increase in DICE score of 0.09 (95% CI of 0.02 to 0.16) for Image ID 1, 0.12 (95% CI of 0.04 to 0.19) for Image ID 2, 0.08 (95% CI of 0.01 to 0.16) for Image ID 3, 0.07 (95% CI of 0.02 to 0.11) for Image ID 5. No evidence of such increase was observed for Image ID 4, with the magnitude of −0.03 (95% CI of −0.07 to 0.01).

**Table 2:** Estimates of DICE change in mixed-effects linear regression.

| Image ID | N | DICE changes, coefficient (95% CI) * | p value |
| --- | --- | --- | --- |
| 1 | 26 | 0.09 (0.02, 0.16) | 0.017 |
| 2 | 27 | 0.12 (0.04, 0.19) | 0.002 |
| 3 | 26 | -0.03 (-0.07, 0.01) | 0.162 |
| 4 | 27 | 0.08 (0.01, 0.16) | 0.043 |
| 5 | 26 | 0.12 (0.05, 0.18) | 0.01 |
| Overall | 132 | 0.07 (0.02, 0.11) | 0.003 |

\*Coefficient estimates the difference of DICE score before and after HDT implementation
adjusting for the random effect of clinicians in mixed effects liner regression model

**Figure 2.**
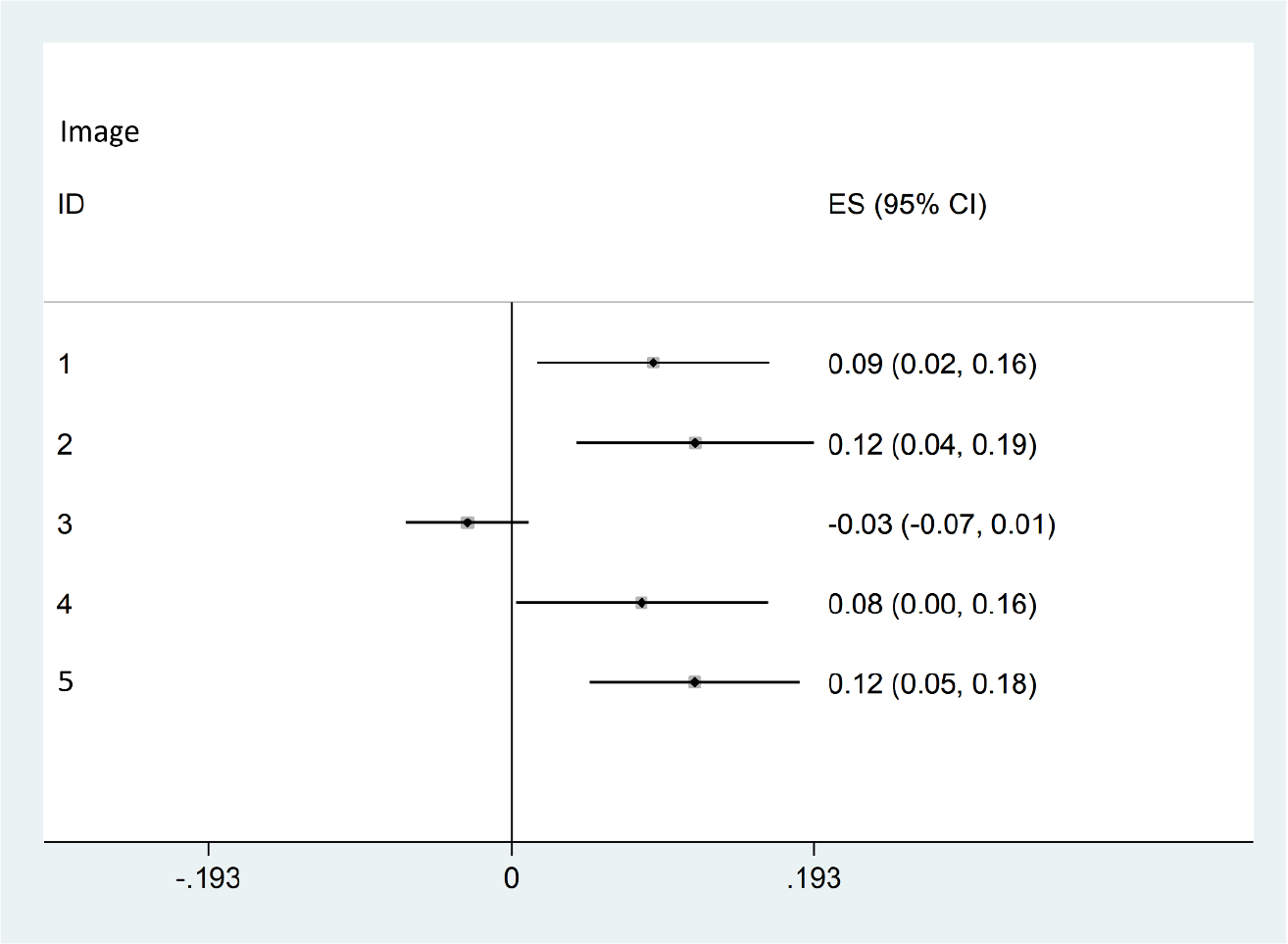
Forest plot of DICE score changes before and after the HDT implementation.

### Performance of clinicians when assessing large versus small lesions

For the slices with large lesions (53 paired segmentations), the mean DICE score was 0.44 (SD 0.31) and 0.48 (SD 0.25) for clinicians before and after the HDT implementation, respectively. No evidence of HDT effect was observed for the large lesion cases with the mean increase in DICE score of 0.05 (95% CI: −0.02, 0.11, p=0.192). For the slices with small lesions (79 paired segmentations), the mean DICE score was 0.25 (SD 0.29) and 0.35 (SD 0.28) for clinicians before and after the HDT implementation, with the respective HDT mean increase in DICE scores following HDT use of 0.08 (95% CI: 0.02, 0.13, p=0.004).

### Performance of consultant versus trainee

Among the 27 clinicians, 12 were consultant neurologists and 15 were trainee neurologists. For the subgroup of 12 consultants the mean DICE score was 0.40 (SD 0.33) and 0.45 (SD of 0.29) before and after the HDT implementation respectively, with no evidence of significant improvement of DICE score for the consultants with the implementation of HDT (mean difference in DICE=0.04, 95% CI: −0.02, 0.107, p=0.189). In contrast, the subgroup of 15 trainees produced a mean DICE score of 0.26 (SD of 0.28) before and 0.37 (SD of 0.26) after the HDT implementation respectively (Figure 2), demonstrating a significant HDT effect in improving DICE scores: mean difference in DICE 0.09 (95% CI: 0.03, 0.14, p=0.004). The performance of consultants versus trainees for each scan is further illustrated in Figure 3.

**Figure 3.**
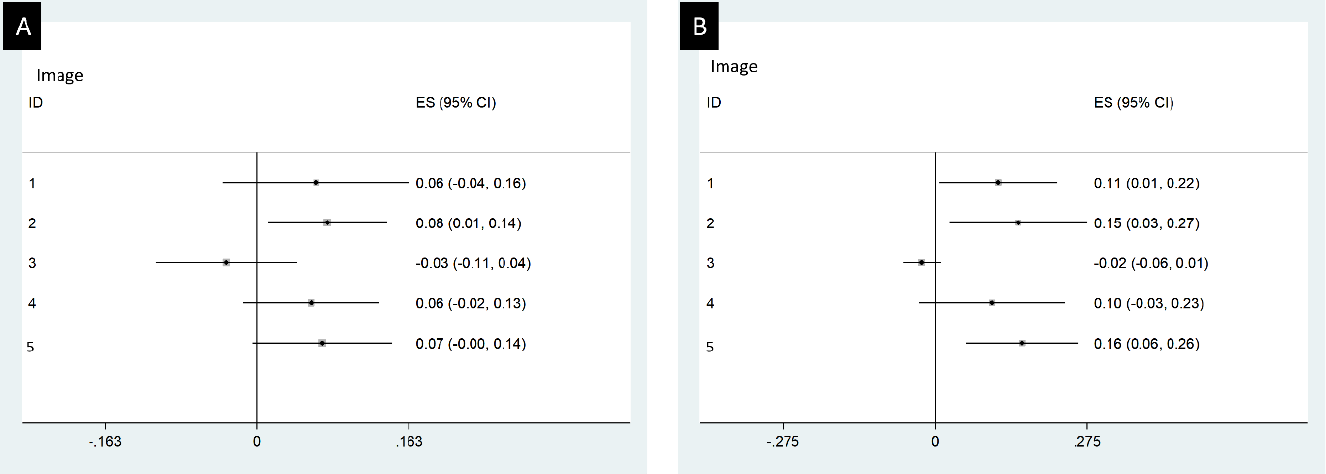
Forest plot of DICE score changes before and after HDT implementation by consultants (A) and trainees (B)

### Success rate of HDT implementation

On review of the data, the clinicians decided not to change their original segmentations after reviewing the HDT output 50% (95% CI: 42%, 58%) of the time. When we analysed only the segmentations that were changed after reviewing the HDT output, we note the following changes to the original segmentation: mean DICE score was 0.42 (SD 0.31) and 0.51 (SD 0.19) respectively, and mean difference of DICE was 0.09 (95%CI: −0.01, 0.19) for the consultants; for the trainees, mean DICE was 0.31 (SD 0.29) and 0.51(SD 0.16) respectively, and difference of mean DICE was 0.20 [0.12, 0.28]).

## Discussion

This Hypodensity Detection Tool (HDT) had relatively good performance in the detection of acute hypodensity on NCCT. It proved to be superior to all clinicians and indeed, overall clinician performance improved with its use. It was most useful for neurology trainees, and when assessing small hypodense lesions. If all neurology trainees had used the HDT, then the results may have been even more impressive.

These results suggest that applying the use of HDT to acute stroke CT would have the greatest impact in regional, rural, and remote health care settings, where expert neurology consultants are not immediately available to assess acute stroke imaging for patients who need reperfusion therapy. Whilst any automated imaging tool does not replace the need for clinical judgment, this HDT could provide support and guidance, and/or notify the onsite doctors (without expertise in NCCT assessment) that the patient being assessed may have hypodensity on their NCCT, which can then help to influence treatment decisions.

One of the most interesting findings of this study was the ‘resistance’ of the doctors to change their assessment of the NCCT, even after reviewing the HDT output. Fifty percent of the doctors were not willing to trust the hypodensity detection tool. This either speaks to the doctor’s (over)confidence in their ability to assess NCCT, or their wariness of relying on HDT to make an imaging decision (which has flow on effects of treatment decisions).

The main limitation of this study was the number of clinicians recruited and the resultant total of paired segmentations. The variability of results comes from the clinicians, with the scans used to test the clinician’s ability. The scans were selected to reflect different characteristics of lesions (size, location) to ensure the representative sample of scans, making them generalisable in acute stroke.

In summary, these findings underscore the potential of HDT to significantly enhance diagnostic precision and hold promise for its valuable application in clinical practice, particularly in neurology training and challenging diagnostic scenarios involving smaller hypodense lesions.

## Data Availability

The authors confirm that the raw data supporting the findings of this study are available from the corresponding author upon reasonable request

## Acknowledgements

The participants of this study; Balabanski A, Sharobeam A, Park A, Lim B, Colman B, Giarola B, Williams C, Esperon C, Murtimer C, Blair C, Preml D, Hg F, Zhao H, Beharry J, Broadley J, Thomas J, Shipley J, Ng J, Wong J, Ja J, Baskin J, Ma M, Linger M, Megans M, Valente M, Nyugen M, Lizark N, Swarup O, Siriratnam P, Holper S, Yogendrakumar V, and Stanislau V.

## Sources of funding

Angela Dos Santos is supported by the Australian Commonwealth Government on a Research Training Program Scholarship.

## Disclosures

Nil

## Notes

### Competing Interest Statement

The authors have declared no competing interest.

### Clinical Trial

NA

### Author Declarations

The INSPIRE had central ethics approval by the Hunter New England Human Research Ethics Committee (HNEHREC 11/08/17/14.01)

## References

1. Bivard A CL, Parsons, M. Artificial intelligence for decision support in acute stroke — current roles and potential. Nat Rev Neurol. 2020;16:575–85.

2. Peter R KP, Blezek D, Oscar Beitia A, Stepan-Buksakowska I, Horinek D, Flemming KD, Erickson BJ. A quantitative symmetry-based analysis of hyperacute ischemic stroke lesions in noncontrast computed tomography. Med Phys. 2017;44(1):192–9.

3. Gao J PM, Kawano H, Levi CR, Evans TJ, Lin L, Bivard A.. Visibility of CT Early Ischemic Change Is Significantly Associated with Time from Stroke Onset to Baseline Scan beyond the First 3 Hours of Stroke Onset. J Stroke. 2017;19(3):340–6.

4. Jl S. Time is brain--quantified. Stroke. 2006;37(1):263–6.

5. RANZCR. Clinical Radiology Workforce Census Report: Australia. 2016.

6. Qiu W KH, Teleg E, Ospel JM, Sohn SI, Almekhlafi M, Goyal M, Hill MD, Demchuk AM, Menon BK. Machine Learning for Detecting Early Infarction in Acute Stroke with Non-Contrast-enhanced CT. Radiology. 2020;294(3):638–44.

7. Przelaskowski A SK, Bargieł P, Walecki J, Biesiadko-Matus-zewska M, Kazubek M. Improved early stroke detection: wavelet-based perception enhancement of computerized tomography exams. Comput Biol Med. 2007;37:524–33.

8. Takahashi N LY, Tsai D-Y, et al. Improvement of detection of hypoattenuation in acute ischemic stroke in unenhanced computed tomography using an adaptive smoothing filter. Acta Radiologica. 2008;49:816–26.

9. Takahashi N TD-Y, Lee Y, Kinoshita T, Ishii K. Z-score mapping method for extracting hypoattenuation areas of hyperacute stroke in unenhanced CT. Acad Radiol. 2010;17:84–92.

10. Gillebert CR HG, Mantini D. Automated delineation of stroke lesions using brain CT images. NeuroImage Clin. 2014;4:540–8.

11. Takahashi N TD-Y, Lee Y, et al. Usefulness of z-score mapping for quantification of extent of hypoattenuation regions of hyperacute stroke in unenhanced computed tomography: analysis of radiologists’ performance. J Comput Assist Tomogr. 2010;34:751–6.

12. Takahashi N LY, Tsai D-Y, Kinoshita T, Ouchi N, Ishii K. Computer-aided detection scheme for identification of hypoattenuation of acute stroke in unenhanced CT. Radiol Phys Technol. 2012;5:98–104.

13. Tang F-H ND, Chow DHK. An image feature approach for computer-aided detection of ischemic stroke. Comput Biol Med. 2011;41:529–36.

14. Nowinski WL GV, Qian G, et al. Automatic detection, localization, and volume estimation of ischemic infarcts in noncontrast computed tomographic scans: method and preliminary results. Radiol. 2013;48:661–70.

15. Avants BB TN, Song G, Cook PA, Klein A, Gee JC. A reproducible evaluation of ANTs similarity metric performance in brain image registration. Neuroimage. 2011;54(3):2033–44.

